# Investigating the relationship of serum vitamin D levels with blood pressure and hypertension risk in The HUNT Study: Using traditional observational and Mendelian randomization approaches

**DOI:** 10.1101/2024.02.13.24302800

**Authors:** Lin Jiang, Yi-Qian Sun, Marion Denos, Ben Michael Brumpton, Yue Chen, Vegard Malmo, Eleanor Sanderson, Xiao-Mei Mai

## Abstract

**Background:** Limited studies have triangulated the relationship between serum vitamin D [25(OH)D] levels and systolic blood pressure (SBP), diastolic blood pressure (DBP) or hypertension risk using traditional observational and Mendelian randomization (MR) approaches.

**Methods and results:** Data were obtained from the Norwegian Trøndelag Health Study (HUNT). A cross-sectional study was performed among 5854 participants from HUNT2. Among them, 3592 participants were followed over 11 years for a prospective analysis. Furthermore, a one-sample MR was conducted with 86,324 participants from HUNT. An externally weighted genetic risk score based on 19 genetic variants for 25(OH)D was used as instrument and the Wald ratio method was applied to evaluate causal associations. Additionally, two-sample MR were performed using updated publicly available data. Our cross-sectional analyses showed a 25 nmol/L increase in 25(OH)D was associated with a 1.73 mmHg decrease in SBP (95 % CI -2.46 to -1.01), a 0.91 mmHg decrease in DBP (95% CI - 1.35 to -0.47) and 19% lower prevalence of hypertension (OR 0.81, 95% CI 0.74 to 0.90) after adjusting for important confounders. However, these associations disappeared in prospective analyses. Both one-sample and two-sample MR results suggested no causal associations.

**Conclusions:** Cross-sectional findings of inverse associations between serum 25(OH)D levels and blood pressure or hypertension were not supported by results from the prospective and MR analyses, suggesting no causal links.

**Clinical Perspective What Is New?:**

- We triangulated the potential relationships of serum 25(OH)D with blood pressure and hypertension using several observational methods such as cross-sectional, prospective cohort, one-sample and two-sample Mendelian randomization (MR) approaches.

**What Are the Clinical Implications?:**

- The consistency across the prospective, one-sample MR and two-sample MR analyses enhanced the robustness of the findings of no causal association between vitamin D and blood pressure or hypertension.
- Clinicians should be cautious when recommending vitamin D supplementation to the general population for the prevention of cardiovascular diseases.

## Introduction

Hypertension is a major risk factor for cardiovascular disease and all-cause mortality that affects approximately one-third of the adult population globally ^1^. Genetic, environmental and lifestyle factors all contribute to the development of hypertension ^1^. Vitamin D is a micronutrient for the body that is synthesized in the skin upon exposure to sunlight or obtained from dietary sources such as fatty fish and from supplements such as cod liver oil ^2^. Vitamin D insufficiency, typically assessed by circulating levels of 25-hydroxyvitamin D [25(OH)D] below 50 nmol/L affects over 50% of the world population, particularly during the winter ^3^. Vitamin D insufficiency has been suggested as a potential modifiable risk factor for hypertension since vitamin D is involved in various physiological processes, including regulation of the renin-angiotensin-aldosterone system, insulin secretion, endothelial function and inflammation ^4,5^.

Most observational studies have reported an inverse association between serum 25(OH)D and blood pressure or hypertension risk ^6^. However, no clear evidence on causality has been found based on Randomized Controlled Trials (RCTs) ^6^. Residual confounding or reverse causality can bias the results in traditional observational studies, while existing RCTs for the topic have limitations such as small sample size, short duration and the inclusion of participants who either had vitamin D deficiency or were older age ^6,7^.

Mendelian randomization (MR) approach uses genetic variants as instrumental variables for the risk factor of interest and can estimate causal effects in the presence of unobserved confounding of the exposure and the outcome ^8^. The advantage of MR is that genetic variants are randomly assigned at conception and remain stable over the lifetime ^9^. Bias due to reverse causation may be avoided and the influence of residual confounding is reduced ^9^. Thus, MR studies can offer supplementary evidence for causal relationships, while being less expensive and time-consuming compared to RCTs.

There is a limited number of MR studies on the relationship of serum 25(OH)D with blood pressure or risk of hypertension ^10–13^. Vimaleswaran et al. reported inverse causal links between genetically determined serum 25(OH)D and diastolic blood pressure (DBP) as well as risk of hypertension, but not systolic blood pressure (SBP) in 146,581 European adults ^12^. They utilized two single nucleotide polymorphisms (SNPs) that were associated with the vitamin D synthesis as instruments. However, recent MR studies using more SNPs in larger sample sizes of over 320,000 participants from the UK Biobank found no causal associations ^10,11^. Furthermore, another MR study suggested that causal associations might be non-linear since they only existed in a subgroup of people with vitamin D deficiency ^13^. Thus, evidence for causality of the associations is inconsistent.

In the current study, we aimed to investigate the potential causal associations between serum 25(OH)D and SBP, DBP or risk of hypertension in the Norwegian Trøndelag Health (HUNT) population using both conventional observational and MR approaches. In addition, we triangulated our results with two-sample MR analyses using publicly available summary data from the latest genome-wide association studies (GWASs). We also explored the potential non-linear causality in the HUNT population.

## Methods

### Study design and population

The HUNT Study is a large population-based health study that has been carried out in four phases, HUNT1 (1984-86), HUNT2 (1995–97), HUNT3 (2006-2008) and HUNT4 (2017-2019) in the Trøndelag county of Norway ^14,15^. All adults aged 20 years or older were invited to complete general questionnaires on health and lifestyle status and undergo clinical examinations ^14,15^.

In total, 96,436 adults participated in HUNT2, HUNT3 or HUNT4 ^14^. We first excluded 10,001 participants without information on genetic variants (Figure 1). Afterwards, 111 participants with missing information on SBP or DBP were excluded, leaving 86,324 adults in the total cohort for the current study. Of the total cohort, a 10% random sample of the HUNT2 population had information on serum 25(OH)D measurements (n=5854), which was regarded as a sub-cohort.

**Figure 1.**
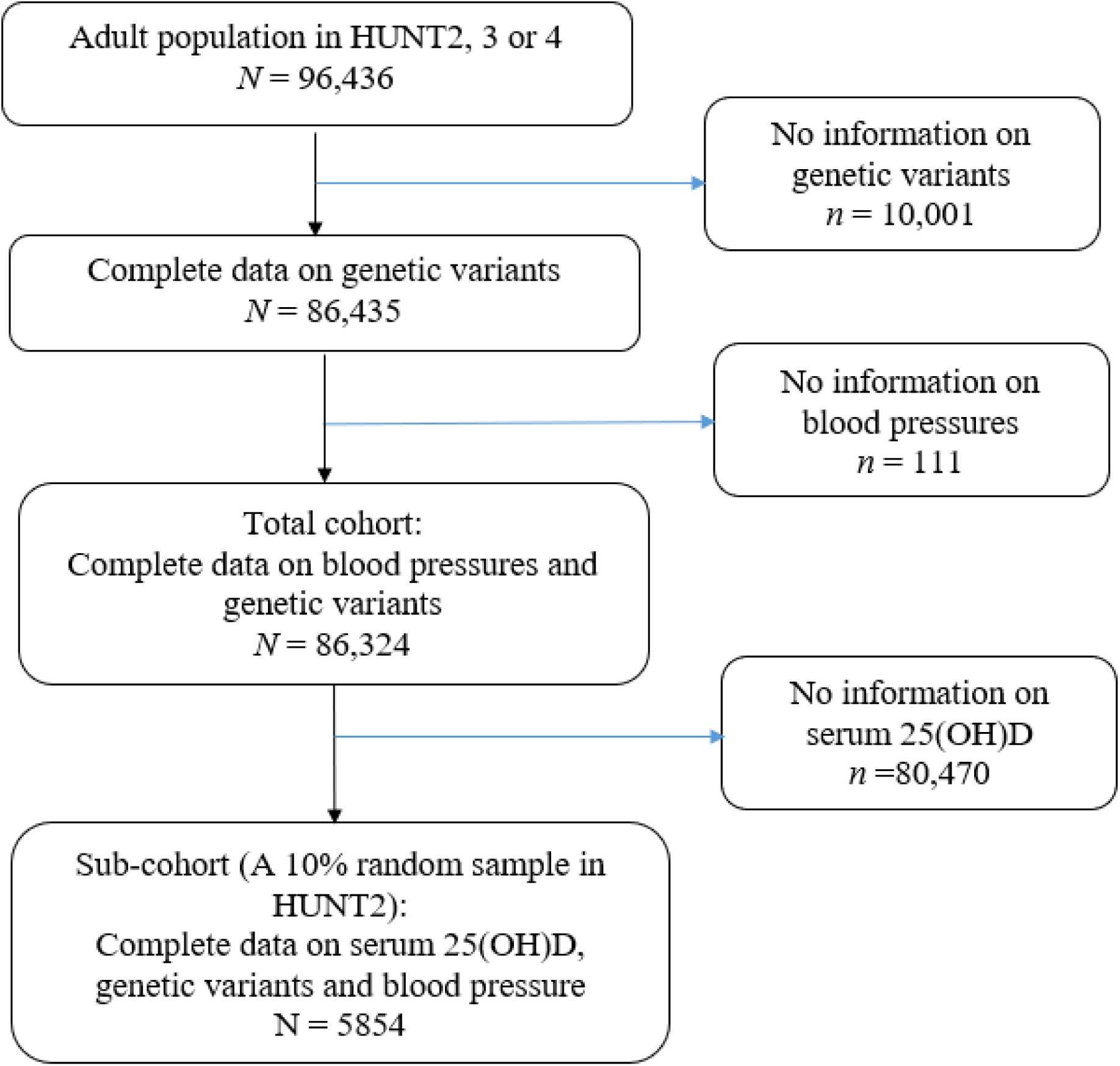
Flow chart of the study population.

### Measurements and standardization of serum 25(OH)D levels

Serum 25(OH)D levels in the sub-cohort were measured at HUNT Biobank using LIAISON 25-OH Vitamin D TOTAL (DiaSorin, Saluggia, Italy), a fully automated, antibody based, chemiluminescence assay. The detection range of the assay for total serum 25(OH)D is 10– 375 nmol/L. Measurements of serum 25(OH)D were seasonally standardized ^16^. This standardized 25(OH)D represents the annual average value of serum 25(OH)D for each participant. In this way, the seasonal fluctuation of the levels owing to the high latitude of Norway could be properly corrected. The seasonally standardized serum 25(OH)D levels were treated as both a continuous variable (per 25 nmol/L increase) and a categorical variable of four categories (<30.0, 30.0−49.9, 50.0−74.9 and ≥75.0 mmol/L) ^17^.

### Vitamin D SNPs and genetic risk score as the instrumental variable

DNA was extracted from blood samples that were collected in HUNT2, HUNT3 or HUNT4 and stored in the HUNT Biobank. Genome-wide genotyping and imputation were carried out with sample and variant quality control by using Illumina Humina HumanCoreExome arrays^18^. We utilized 21 SNPs derived from four gene regions (GC, DHCR7, CYP2R1 and CYP24A1) as candidate instrument variables for serum 25(OH)D, reported by Sofianopoulou et al. ^19^. These SNPs were proven to have strong associations with serum 25(OH)D (P value <5 x 10^-8^) and selected based on their clear biological roles in vitamin D transport, synthesis and metabolism ^19^.

Information on 2 SNPs (rs139148694 and rs35870583) was missing in the HUNT data since they did not pass imputation quality control (R^2^ of linkage disequilibrium >0.8), leaving 19 SNPs to construct an externally weighted genetic risk score (GRS) for our analyses. Using a GRS instead of the individual genetic variants can ensure that a large proportion of serum 25(OH)D can be accounted for and therefore reduce weak instrument bias and increase statistical power ^20^. The externally weighted GRS was calculated as the sum of the number of effect alleles carried for each SNP weighted by the reported beta coefficient (β) for serum 25(OH)D derived from the study by Sofianopoulou et al. ^19^. Compared to other GRSs with SNPs selected either biologically driven or statistically driven ^13,21–23^, the GRS composed of the 19 SNPs, selected based on a biologically driven approach ^19^, emerged as a more robust instrument for serum 25(OH)D in the HUNT dataset (Table 1). This GRS explained 5.6% of the variability in serum 25(OH)D among the HUNT population with a F-statistic of 348. The characteristics of the 19 individual SNPs are shown in Supplementary Table 1.

**Table 1.** Comparing instrument strength of GRS**s** constructed with SNPs selected via different methods in association with serum 25(OH)D levels measured in the HUNT^1^ Study.

| Methods to select instruments | Original papers | Original population (N) | Numbers of SNPs<sup>2</sup> | First-stage F statistics | R<sup>2</sup> value |
| --- | --- | --- | --- | --- | --- |
| Biologically driven | Sofianopoulou et al. [1] | UK Biobank+EPIIC-CVD (N=355,144) | 19 | 348 | 0.056 |
| Biologically driven | Jiang et al. [2] | SUNLIGHT Consortium (N=79,366) | 6 | 225 | 0.037 |
| Statistically driven | Zhou et al. [3]<sup>3</sup> | UK Biobank, replicated in SUNLIGHT (N =294,970) | 33 | 255 | 0.042 |
| Statistically driven | Manousaki et al. [4] | UK Biobank+SUNLIGHT (N=443,734) | 60 | 294 | 0.049 |

EPIIC-CVD: the European Prospective Investigation into Cancer and Nutrition Cardiovascular Disease study; GRS: Genetic risk score; GWAS: Genome-wide association study; HUNT: The Trøndelag Health Study; SNP: Single Nucleotide Polymorphisms; SUNLIGHT: Study of Underlying Genetic Determinants of Vitamin D and Highly Related Traits; 25(OH)D: 25-hydroxyvitamin D
<sup>1</sup> HUNT: The HUNT population used for testing instrument strength was based on a 10% random sample with complete information on serum 25(OH)D, genetic variants and blood pressure measurements from HUNT2 (n=5854).
<sup>2</sup> Two SNPs (rs139148694 and rs35870583) from Sofianopoulou et al., 2 SNPs (rs7522116 and rs12798050) from Zhou et al., and 9 SNPs (rs2934744, rs144613541, rs11127048, rs7650253, rs7699711, rs3822868, rs9668081, rs71383766, rs112285002) from Manousaki et al. were not available in HUNT.
<sup>3</sup> The original GWAS was by Revez et al. 2020 [5] using UK Biobank data (N=417,580). In the study by Zhou et al., the authors constructed a weighted genetic score based on 35 SNPs detected in the GWAS by Revez et al. and replicated in the SUNLIGHT Consortium data.

### Other baseline variables

In HUNT2, HUNT3 and HUNT4, body weight and height were measured by health professionals at clinical examination. Height was measured to the nearest centimeters and weight to the nearest 0.5 kg. Body mass index (BMI) was calculated as weight in kilograms divided by height squared in meter (kg/m^2^). Other covariates were categorized as: sex (women and men), smoking status with detailed information on pack-years (pyrs) [(never, former (<10, 10-20 and >20 pyrs) and current (<10, 10-20 and >20 pyrs)], alcohol consumption (never, 1–4 and ≥5 times/month), leisure physical activity (inactive, low, moderate and active). Missing information on each of the mentioned variables was included in the analyses as an “unknown” category. For adults who participated in more than one HUNT survey, data were retrieved from the first HUNT measurement if available except for BMI. We used the mean value of BMI for those participating in at least two surveys. Information on education (<10, 10-12 and ≥13 years) and economic difficulty (yes and no) was collected in HUNT2 only. We used the same categorization of variables as did in previous HUNT publications ^24,25^. Batch for genotyping and 20 principal components (PCs) of ancestry were included as covariates in the MR analyses.

### Measurements of blood pressure and hypertension

In HUNT2, HUNT3 and HUNT4, SBP and DBP were measured three times by trained nurses using an automatic oscillometry (Dinamap, Critikon, Florida) with 1-min interval after the participants had rested for several minutes in a sitting position. Cuff size was adjusted according to arm circumference. The mean value of the last two measurements were used in the current study. In the total cohort, data on SBP and DBP were retrieved from the first HUNT measurement for adults who participated in at least two HUNT surveys. To account for bias due to use of antihypertensive medication, blood pressure measurements were, based on recommendations by Cui et al. ^26^ and Tobin et al. ^27^, amended by adding 10 and 5 mmHg to the measured SBP and DBP among participants who self-reported to use antihypertensive medication in each HUNT survey, respectively. Hypertension was defined as SBP ≥140 mmHg or DBP ≥90 mmHg or self-reported use of anti-hypertensive medication in each survey ^12,28,29^.

### Statistical analyses

First, we performed cross-sectional analyses on the assocations between serum 25(OH)D and SBP, DBP and hypertension in the sub-cohort of the HUNT2 population (n=5854). Second, we conducted prospective analyses among the participants in the sub-cohort who were followed up from HUNT2 to HUNT3 for an average 11 years (n=3592). Linear regression was used to examine the associations between serum 25(OH)D and SBP or DBP, while logistic regression was used for the association with hypertension. Potential confounders in the cross-sectional and prospective analyses were age at baseline, sex, BMI, smoking status with pack-years, alcohol consumption, leisure physical activity, education and social economy difficulty based on previous knowledge ^30–32^. For the prospective associations of serum 25(OH)D in HUNT2 with SBP and DBP in HUNT3 among the 3592 participants, baseline SBP and DBP in HUNT2 were additionally adjusted for. For the prospective association with the risk of hypertension, the analyses were performed among 2227 of the 3592 participants who did not have hypertension in HUNT2.

Third, we performed a one-sample MR study in which the GRS−outcome association was assessed in the total cohort while the GRS−exposure association was assessed in the sub-cohort ^33^. A Wald ratio method was applied to compute the MR estimates ^8,34^. We calculated the MR estimate as a ratio of the coefficient of the GRS−outcome (SBP, DBP or hypertension) association over the coefficient of GRS−exposure (Serum 25(OH)D) association ^34,35^. In the case where the outcome was hypertension, we computed a MR-driven odds ratio by applying the natural exponential function of the ratio of coefficients. All regression models were adjusted for age, sex, batch and 20 PCs ^19^.

Three key assumptions should be met for the MR analyses: 1) the GRS should be associated with the serum 25(OH)D levels (relevance assumption); 2) the GRS should not be associated with any potential confounders of the observational associations (independence assumption); and 3) there should be no horizontally pleiotropic effect of the vitamin D SNPs on SBP, DBP or hypertension risk (exclusion assumption). We tested the first assumption in the sub-cohort using a F statistic and R^2^ value for the association between GRS and serum 25(OH)D. The GRS is considered to be adequate instrument variable if F-statistic >10 ^36^. To address the second assumption, we tested the associations between the GRS and the available confounders in the sub-cohort using linear or logistic regression. We assessed the third assumption using SNP-based two-sample methods such as MR-Egger ^37^, weighted median ^38^ and MR-Pleiotropy Residual Sum and Outlier (MR-PRESSO) ^39^ methods (Supplementary text 1).

Since we couldn’t obtain summary data for the 19 SNPs from Sofianopoulou et al. ^19^, we utilized three other sets of SNPs as instruments for the two-sample MR analyses to test our findings ^13,21,23^ (Supplementary Table 4 and Supplementary text 2). The first set, from the SUNLIGHT Consortium by Jiang et al. (N=79,366), contained 6 SNPs ^21^. We also utilized SNP sets from the recent GWASs, including 35 SNPs from Zhou et al. (N=294,770) ^13^ and 69 from Manousaki et al. (N=443,734) ^23^. For outcome data, the largest GWAS for SBP and DBP by Evengelou et al. using the UK Biobank data (N=757,601) ^40^ and GWAS from FinnGen comprising 42,857 hypertension cases and 218,792 controls ^41^ were used. The set of 6 SNPs was chosen as our primary instruments for the two-sample MR analyses due to their clear biological relevance to serum 25(OH)D and the absence of overlap between the exposure and outcome GWAS datasets.

In addition, we briefly tested if there existed non-linear causal associations using both the residual method ^19^ and the doubly-ranked method ^42^ among the HUNT sub-cohort (Supplementary text 3). The residual method divides the population into equal-sized strata using exposure residuals ^19,43^, assuming constant genetic effect on the exposure within each stratum. The doubly-ranked method is a non-parametric stratification method which is less sensitive to this assumption ^42^.

All statistical analyses were performed with STATA/SE 16.1 (College Station, TX, USA) or R (4.0.2). The package “TwoSampleMR” was used for the two-sample MR and package “SUMnlmr” for non-linear MR in R.

## Results

The baseline characteristics of the participants in the total cohort and sub-cohort showed slightly different distributions (Table 2). Participants in the sub-cohort were older, had higher average SBP and DBP, and were more likely to be current smokers and non-drinkers compared to the total cohort.

**Table 2.**
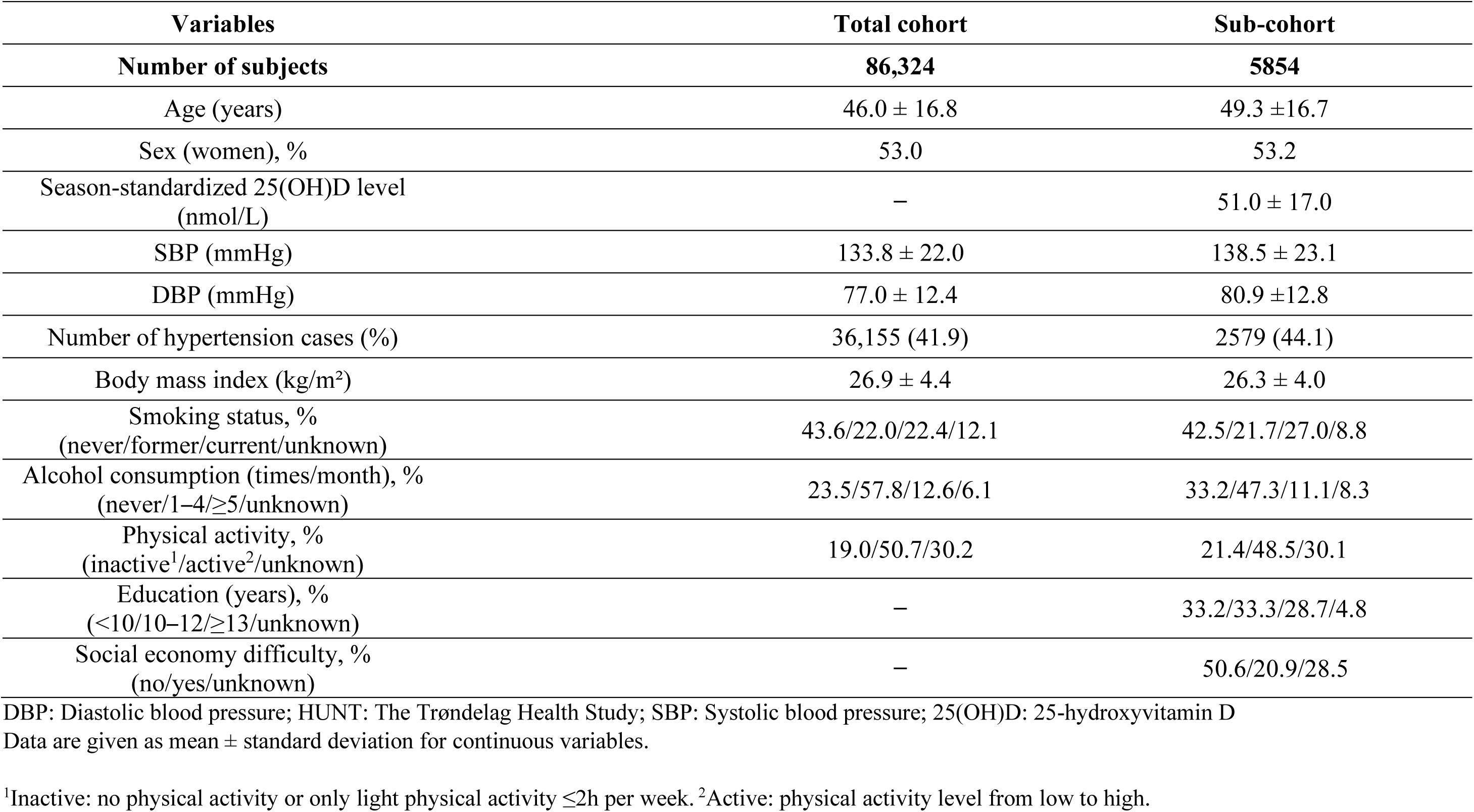
Baseline characteristics of participants in the analysis total cohort and sub-cohort of the HUNT Study.

| Variables | Total cohort | Sub-cohort |
| --- | --- | --- |
| **Number of subjects** | **86,324** | **5854** |
| Age (years) | 46.0 ± 16.8 | 49.3 ± 16.7 |
| Sex (women), % | 53.0 | 53.2 |
| Season-standardized 25(OH)D level (nmol/L) | – | 51.0 ± 17.0 |
| SBP (mmHg) | 133.8 ± 22.0 | 138.5 ± 23.1 |
| DBP (mmHg) | 77.0 ± 12.4 | 80.9 ± 12.8 |
| Number of hypertension cases (%) | 36,155 (41.9) | 2579 (44.1) |
| Body mass index (kg/m<sup>2</sup>) | 26.9 ± 4.4 | 26.3 ± 4.0 |
| Smoking status, %<br>(never/former/current/unknown) | 43.6/22.0/22.4/12.1 | 42.5/21.7/27.0/8.8 |
| Alcohol consumption (times/month), %<br>(never/1–4/≥5/unknown) | 23.5/57.8/12.6/6.1 | 33.2/47.3/11.1/8.3 |
| Physical activity, %<br>(inactive<sup>1</sup>/active<sup>2</sup>/unknown) | 19.0/50.7/30.2 | 21.4/48.5/30.1 |
| Education (years), %<br>(<10/10–12/≥13/unknown) | – | 33.2/33.3/28.7/4.8 |
| Social economy difficulty, %<br>(no/yes/unknown) | – | 50.6/20.9/28.5 |

DBP: Diastolic blood pressure; HUNT: The Trøndelag Health Study; SBP: Systolic blood pressure; 25(OH)D: 25-hydroxyvitamin D
Data are given as mean ± standard deviation for continuous variables.
<sup>1</sup>Inactive: no physical activity or only light physical activity ≤2h per week. <sup>2</sup>Active: physical activity level from low to high.

In the cross-sectional analyses, 5854 participants from HUNT2 in the sub-cohort were included. Among them, 2579 cases of hypertension were identified. We observed that lower serum 25(OH)D were associated with higher SBP and DBP as well as an increased prevalence of hypertension after adjustment for the potential confounders (Table 3, Supplementary Figure 1). Specifically, each 25 nmol/L increase in 25(OH)D was associated with a decrease of 1.73 mmHg in SBP (95 % CI -2.46 to -1.01), a 0.91 mmHg decrease in DBP (95% CI -1.35 to -0.47) and a 19% lower prevalence of hypertension (odds ratio 0.81, 95% CI 0.74 to 0.90). In the prospective analyses, 3592 participants were followed up from HUNT2 to HUNT3. After excluding 1365 participants with hypertension at baseline, 2227 participants were followed until HUNT3 and 542 new diagnosed hypertension cases were found. No associations were found after adjustment for the same confounders (coefficient 0.26 mmHg, 95% CI -0.59 to 1.12 for SBP, coefficient 0.31 mmHg, 95% CI -0.21 to 0.82 for DBP and odds ratio 0.92, 95% CI 0.77 to 1.11 for hypertension for each 25 nmol/L increase in serum 25(OH)D). Results by the categorical variable of serum 25(OH)D were also presented in Table 3.

**Table 3.**
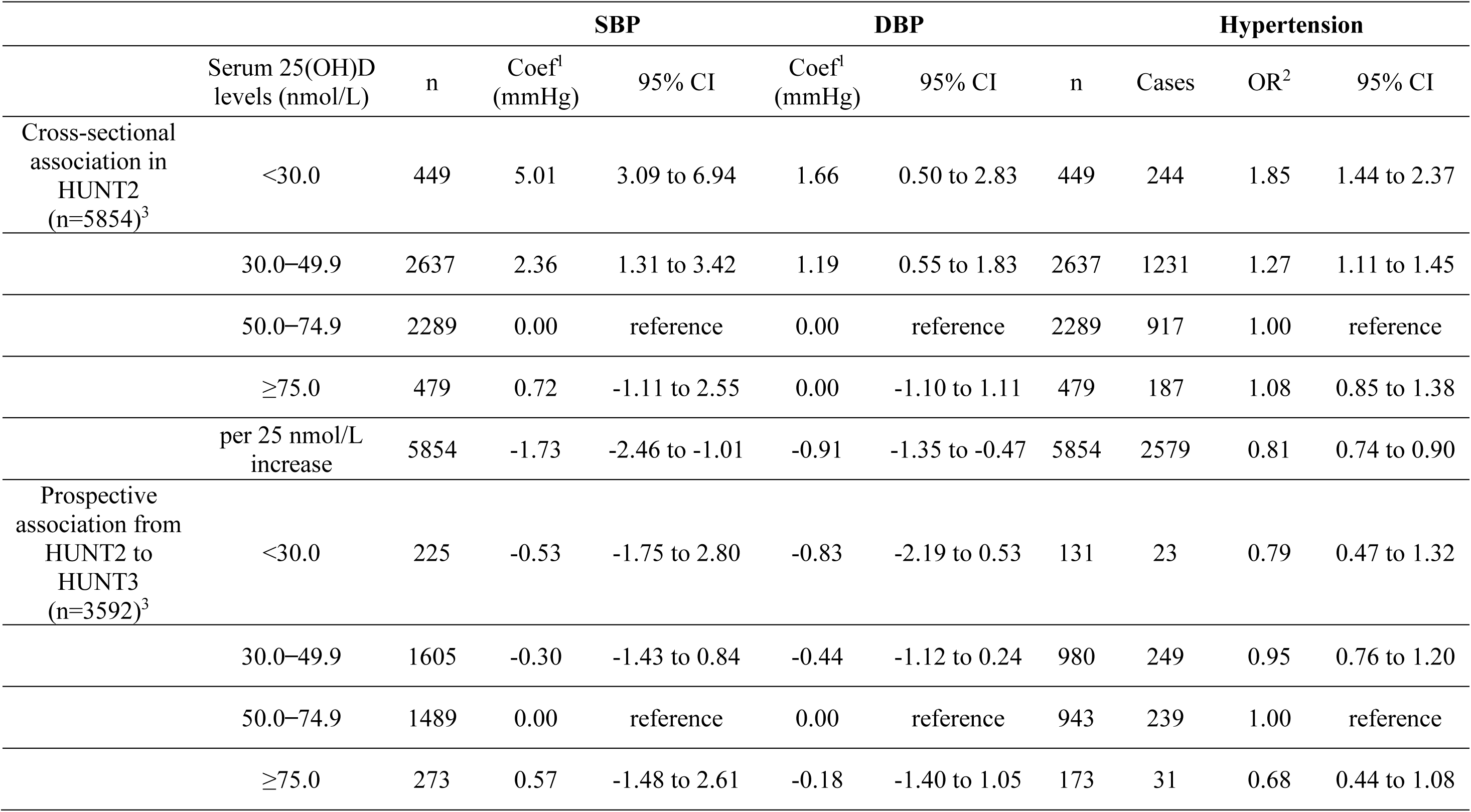
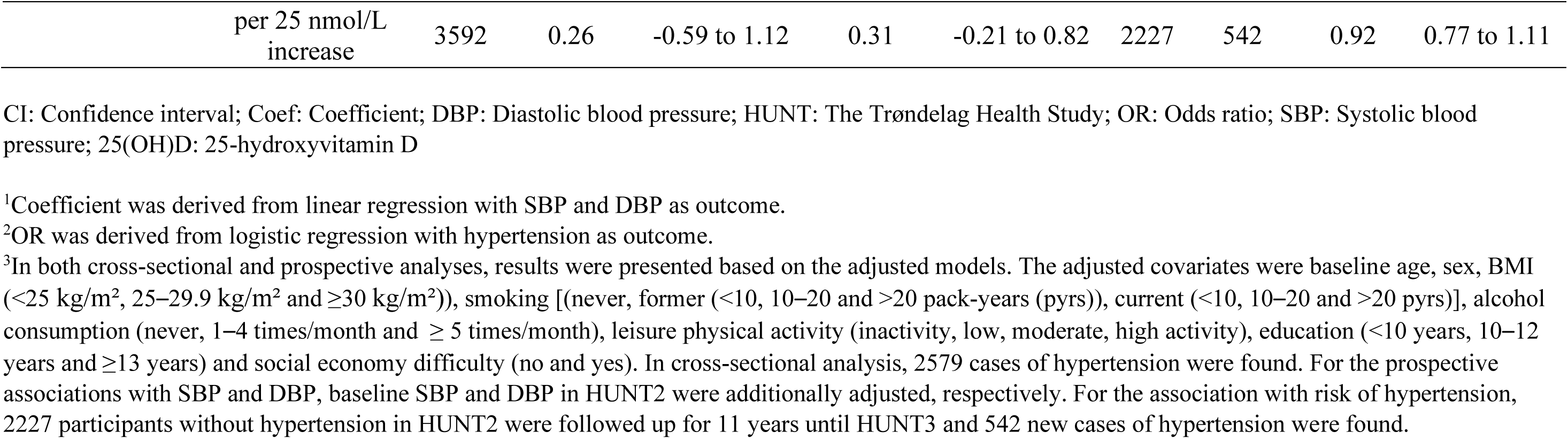
The observational associations of serum 25(OH)D levels with systolic and diastolic blood pressure and hypertension in a sub-cohort of the HUNT2 population.

Results from our one-sample MR analyses showed no causal associations of genetically determined serum 25(OH)D with SBP, DBP and risk of hypertension in the HUNT population (Table 4, Supplementary Figure 1). For each genetically determined 25 nmol/L increase in serum 25 (OH)D, the MR regression coefficient estimate was -0.11 mmHg (95% CI -0.86 to 0.63) for SBP and was 0.04 mmHg (95% CI -0.42 to 0.50) for DBP and the MR odds ratio estimate was 1.04 (95% CI 0.94 to 1.15) for risk of hypertension. The GRS was not associated with the measured confounders in the sub-cohort (Supplementary Table 2). Cochran’s Q tests suggested no heterogeneity (P for Q>0.05, results not shown). Sensitivity analyses using MR-Egger and weighted median methods supported our null findings (Supplementary Table 3, Supplementary Figure 1). The intercepts from MR-Egger method did not deviate markedly from zero and the P values for intercept were all above 0.05. Thus, MR-Egger did not show evidence of a directional pleiotropic effect under the Instrument Strength Independent of Direct Effect assumption. The results based on the MR-PRESSO method suggested no outlier SNPs in the analyses with SBP and DBP. One outlier (rs12794714) was detected with the risk of hypertension. After removing this outlier, the result was similar to our original result for hypertension (Supplementary Table 3).

**Table 4.** One-sample MR^1^ results for the causal associations of serum 25(OH)D levels (per 25 nmol/L increase) with systolic blood pressure, diastolic blood pressure and risk of hypertension in the HUNT Study (N=86,324)

| Outcomes | Coef (mmHg)/OR<sup>2</sup> | 95% CI | P value |
| --- | --- | --- | --- |
| SBP | -0.11 | -0.86 to 0.63 | 0.76 |
| DBP | 0.04 | -0.42 to 0.50 | 0.86 |
| Hypertension | 1.04 | 0.94 to 1.14 | 0.44 |

CI: Confidence interval; Coef: Coefficient; DBP: Diastolic blood pressure; HUNT: The Trøndelag Health Study; MR: Mendelian randomization; OR: Odds ratio; SBP: Systolic blood pressure; 25(OH)D: 25-hydroxyvitamin D
<sup>1</sup>One-sample MR was conducted using the Wald ratio method to compute the MR estimates [6,7]. An externally weighted genetic risk score (GRS) based on 19 serum 25(OH)D-associated genetic variants was used as instrument [1,8]. The MR estimate was obtained by taking the ratio of the coefficient for the GRS-outcome association over the coefficient for GRS-exposure association [6,9]. The coefficients for the GRS-outcome (SBP, DBP and hypertension) were derived within the total cohort (N=86,324). The coefficient for GRS-exposure (serum 25(OH)D levels) was derived within a sub-cohort consisting of 5854 participants. All regression models were adjusted for age, sex, batch and 20 PCs [1].
<sup>2</sup>OR was obtained by applying the natural exponential function of the ratio of the coefficient of GRS-hypertension association over the coefficient of GRS-serum 25(OH)D association.

In the two-sample MR analyses, no causal associations were found between genetically determined serum 25(OH)D and SBP, DBP or risk of hypertension using different sets of SNPs as instruments (Supplementary Tables 5,6, Supplementary Figure 1).

Finally, the P values for testing non-linearity using the residual and doubly-ranked methods were >0.22 for all the three outcomes within the HUNT sub-cohort (results not shown), even though there seemed to be an inverse association between serum 25(OH)D and SBP within the lowest stratum in both methods (Supplementary Table 7).

## Discussion

We observed inverse associations between serum 25(OH)D and SBP, DBP and hypertension in the cross-sectional analyses, but the associations disappeared in the prospective analyses. Our MR analyses provided further evidence supporting the absence of causal associations. The inverse associations of serum 25(OH)D with blood pressure and hypertension observed in our cross-sectional analyses were in line with findings from previous meta-analyses of observational studies ^6,44^. However, the observed inverse associations may not be causal. This is supported by the inconsistent findings between our cross-sectional and prospective analyses and further confirmed by our one-sample and two-sample MR studies. The presence of residual confounding might lead to biased associations. For instance, in observational studies examining the relationship between serum 25(OH)D and cardiovascular disease, BMI often serves as a significant confounder ^45^. Even when adjusted for, traditional observational studies may not fully account for the complexity of adiposity, leaving room for potential residual confounding that introduces bias into the results. On the other hand, individuals with high blood pressure may engage in less outdoor physical activity, leading to reduced sun exposure, and consequently, lower vitamin D synthesis in the body ^6^. This scenario raises the possibility of reverse causation, and it may also be the reason for the observed inverse associations.

MR studies are considered as natural experiments as they utilize genetic variants that are randomly inherited at birth and provide valuable and complementary evidence for causal inference ^9^. In our one-sample MR using data from the HUNT population and two-sample MR using summary data from the latest GWASs, we did not find any causal association between serum 25(OH)D and blood pressure or hypertension. The findings were consistent with three out of four of the published MR studies ^10,11,46^. Only one study showed a marginal decrease in DBP of 0.29 mmHg (95% CI -0.52 to -0.07) and an odds ratio of 0.92 (95% CI 0.87 to 0.97) for hypertension per 10% increase in 25(OH)D level ^12^, using the instrument of a synthesis score based on 2 unweighted SNPs ^47^. This instrument explained only 0.5% variation of the serum 25(OH)D levels with a F statistic value of 220. In our one-sample MR, we constructed an external weighted GRS based on 19 SNPs identified in the UK Biobank population. The 19 SNPs exhibited clear biological relevance in the transport, synthesis and metabolism of serum 25(OH)D ^19^. This GRS explained a significantly greater proportion of the variation (5.6%) of serum 25(OH)D with an F statistic value of 348 in the HUNT population. It showed no obvious directional pleiotropic effects, ensuring its status as a robust genetic instrument.

Using a residual non-linear MR method, Zhou et al. found a non-linear L-shaped association between serum 25(OH)D and blood pressure in the UK Biobank cohort (N=270,000) ^13^. The non-linear association was also suggested in recent meta-analyses of observational studies ^6,44^. Nonetheless, there is a concern that the residual method may violate the assumption of constant genetic effects on serum 25(OH)D within each stratum ^48^. To address this, the doubly-ranked method was developed to provide less biased estimate even in facing the violation of the constant assumption ^42^. However, a recent study suggested that both methods may have yielded biased estimates because the authors found non-null associations between genetic instrument for serum 25(OH)D and age or sex as negative control outcomes within the UK Biobank data, in which the expected results should be null ^48^. Hence, the observed inverse association between serum 25(OH)D and SBP within the lowest stratum using both methods in our study might also be due to unknown biases. Thus, we should be cautious to interpret our results from the non-linear methods.

Our study aimed to triangulate the potential relationships of serum 25(OH)D with blood pressure and hypertension using the HUNT data and summary data from the largest GWASs to date. The consistency across the prospective, one-sample MR and two-sample MR analyses enhanced the robustness of the findings of no causal association between vitamin D and blood pressure outcomes. Using the HUNT data, a large and homogenous population, we had the opportunity to investigate all three MR assumptions. First, the instrument based on the 19 SNPs that we used in the one-sample MR was proven to be a good instrument with adequate strength. Second, we were able to investigate the associations between the GRS and a panel of potential confounders due to the detailed lifestyle and clinical data available in the HUNT dataset. Third, there was no obvious violation of the exclusion assumption since we did not observe horizontal pleiotropy based on the results from the MR-Egger and MR-PRESSO methods.

Our study had several limitations. First, selection bias in the main analyses may exist since participants were healthier than non-participants ^49^. Moreover, there were some differences in baseline characteristics between the total cohort and the sub-cohort. Second, we cannot either confirm or rule out the possibility of non-linear associations since the sample size of our sub-cohort for the non-linear MR analyses was rather small and there are limitations of the current non-linear methods. Third, although HUNT population is a homogeneous population with over 97% Caucasian which minimizes population stratification bias ^50^, it may limit the generalizability of the findings to other ethnic groups.

## Conclusion

Although we observed inverse associations between serum 25(OH)D levels and blood pressure and hypertension in cross-sectional analyses of the Norwegian HUNT population, our results based on prospective analyses, one-sample and two-sample MR suggested a lack of causal associations.

### Abbreviations and symbols

BMI: Body mass index
CI: Confidence interval
DBP: Diastolic blood pressure
GRS: genetic risk score
GWAS: Genome-wide association study
HUNT: The Trøndelag Health Study
MR: Mendelian randomization
MR-PRESSO: MR-Pleiotropy Residual Sum and Outlier
OR: Odds ratio
RCTs: Randomized Controlled Trials
PCs: Principal components
SBP: Systolic blood pressure
SNP: Single Nucleotide Polymorphisms
SUNLIGHT: Study of Underlying Genetic Determinants of Vitamin D and Highly Related Traits
25(OH)D: 25-hydroxyvitamin D

## Data Availability

Data from the HUNT Study is available on request to the HUNT Data Access Committee (hunt@ medisin.ntnu.no) when is used in research projects. The HUNT data access information describes the policy regarding data availability (https://www.ntnu.edu/hunt/data).

## Acknowledgments

The HUNT Study is collaboration between HUNT Research Centre (Faculty of Medicine and Health Sciences, Norwegian University of Science and Technology - NTNU), Nord-Trøndelag County Council, Central Norway Regional Health Authority and the Norwegian Institute of Public Health. The genotyping in HUNT was financed by the National Institutes of Health; University of Michigan; the Research Council of Norway; the Liaison Committee for Education, Research and Innovation in Central Norway; and the Joint Research Committee between St Olavs hospital and the Faculty of Medicine and Health Sciences, NTNU.

## Declarations

### Ethics approval

The study was approved by the Regional Committee for Medical and Health Research Ethics of central Norway (application number:434217). All participants signed informed written consent on participation in HUNT. The study was performed in accordance with the ethical standards as laid down in the 1964 Declaration of Helsinki and its later amendments or comparable ethical standards.

### Consent for participate

Not applicable.

### Consent for publication

Not applicable.

### Authors’ contributions

LJ was responsible for data collection, conducted statistical analyses, interpreted results and wrote the initial draft of the manuscript. YQS and XMM contributed to the study design and statistical analyses. BMB, MD, YQS, YC, VM, ES and XMM participated in the data interpretation, contributed to the manuscript writing with important intellectual content. All authors approved the final version of the manuscript.

## References

1. Mills KT, Stefanescu A, He J. The global epidemiology of hypertension. **Nat Rev Nephrol**. 2020;16(4):223–237.

2. Basit S. Vitamin D in health and disease: a literature review. **Br J Biomed Sci**. 2013/01/01 2013;70(4):161-172. doi:10.1080/09674845.2013.11669951

3. van Schoor NM, Lips P. Worldwide vitamin D status. **Best Pract Res Clin Endocrinol Metab**. Aug 2011;25(4):671–80. doi:10.1016/j.beem.2011.06.007

4. Azeim NAHA, Ragab HM, Shaheen HMEM, Awad SMB. An Overview About Vitamin D Role in Human Health. **J Pharm Negat Results**. 2023:373–377.

5. Kunutsor SK, Apekey TA, Steur M. Vitamin D and risk of future hypertension: meta-analysis of 283,537 participants. **Eur J Epidemiol**. 2013;28:205–221.

6. Zhang D. Effect of vitamin D on blood pressure and hypertension in the general population: an update meta-analysis of cohort studies and randomized controlled trials. **Prev Chronic Dis**. 2020;17:E03

7. Bouillon R, Manousaki D, Rosen C, Trajanoska K, Rivadeneira F, Richards JB. The health effects of vitamin D supplementation: evidence from human studies. **Nat Rev Endocrinol**. Feb 2022;18(2):96–110. doi:10.1038/s41574-021-00593-z

8. Sanderson E, Glymour MM, Holmes MV, et al. Mendelian randomization. **Nat Rev Methods Primers**. 2022/02/10 2022;2(1):6. doi:10.1038/s43586-021-00092-5

9. Lawlor DA, Harbord RM, Sterne JA, Timpson N, Davey Smith G. Mendelian randomization: using genes as instruments for making causal inferences in epidemiology. **Stat Med**. 2008;27(8):1133–1163.

10. Meng X, Li X, Timofeeva MN, et al. Phenome-wide Mendelian-randomization study of genetically determined vitamin D on multiple health outcomes using the UK Biobank study. **Int J Epidemiol**. Oct 1 2019;48(5):1425–1434. doi:10.1093/ije/dyz182

11. Giontella A, Lotta LA, Baras A, et al. Calcium, Its Regulatory Hormones, and Their Causal Role on Blood Pressure: A Two-Sample Mendelian Randomization Study. **J Clinical Endocrinol Metab**. 2022;107(11):3080–3085.

12. Vimaleswaran KS, Cavadino A, Berry DJ, et al. Association of vitamin D status with arterial blood pressure and hypertension risk: a mendelian randomisation study. **Lancet Diabetes Endocrinol**. Sep 2014;2(9):719–29. doi:10.1016/s2213-8587(14)70113-5

13. Zhou A, Selvanayagam JB, Hyppönen E. Non-linear Mendelian randomization analyses support a role for vitamin D deficiency in cardiovascular disease risk. **Eur Heart J**. 2022;7;43(18):1731-1739doi:10.1093/eurheartj/ehab809

14. Krokstad S, Langhammer A, Hveem K, et al. Cohort Profile: the HUNT Study, Norway. **Int J Epidemiol**. Aug 2013;42(4):968–77. doi:10.1093/ije/dys095

15. Åsvold BO, Langhammer A, Rehn TA, et al. Cohort Profile Update: The HUNT Study, Norway. **Int J Epidemiol**. Feb 8 2023;52(1):e80–e91. doi:10.1093/ije/dyac095

16. Degerud E, Hoff R, Nygård O, et al. Cosinor modelling of seasonal variation in 25-hydroxyvitamin D concentrations in cardiovascular patients in Norway. **Eur J Clin Nutr**. 2016/04/01 2016;70(4):517-522. doi:10.1038/ejcn.2015.200

17. Institute of Medicine Committee to Review Dietary Reference Intakes for Vitamin D, Calcium. The National Academies Collection: Reports funded by National Institutes of Health. In: Ross AC, Taylor CL, Yaktine AL, Del Valle HB, eds. *Dietary Reference Intakes for Calcium and Vitamin D*. National Academies Press (US) Copyright © 2011, National Academy of Sciences.; 2011.

18. Ferreira MA, Vonk JM, Baurecht H, et al. Shared genetic origin of asthma, hay fever and eczema elucidates allergic disease biology. **Nat genet**. 2017;49(12):1752–1757.

19. Sofianopoulou E KS, Afzal S, Jiang T, Gill D, Gundersen TE, et al. Estimating dose-response relationships for vitamin D with coronary heart disease, stroke, and all-cause mortality: observational and Mendelian randomisation analyses. **Lancet Diabetes Endocrinol**. Dec 2021;9(12):837–846. doi:10.1016/s2213-8587(21)00263-1

20. Burgess S, Thompson SG. Use of allele scores as instrumental variables for Mendelian randomization. **Int J Epidemiol**. 2013;42(4):1134–1144.

21. Jiang X, O’Reilly PF, Aschard H, et al. Genome-wide association study in 79,366 European-ancestry individuals informs the genetic architecture of 25-hydroxyvitamin D levels. **Nat Commun**. Jan 17 2018;9(1):260. doi:10.1038/s41467-017-02662-2

22. Revez JA, Lin T, Qiao Z, et al. Genome-wide association study identifies 143 loci associated with 25 hydroxyvitamin D concentration. **Nat Commun**. 2020/04/02 2020;11(1):1647. doi:10.1038/s41467-020-15421-7

23. Manousaki D, Mitchell R, Dudding T, et al. Genome-wide Association Study for Vitamin D Levels Reveals 69 Independent Loci. **Am J Hum Genet**. Mar 5 2020;106(3):327–337. doi:10.1016/j.ajhg.2020.01.017

24. Jiang L, Sun Y-Q, Brumpton BM, Langhammer A, Chen Y, Mai X-M. Body mass index and incidence of lung cancer in the HUNT study: using observational and Mendelian randomization approaches. **BMC Cancer**. 2022;22(1):1152.

25. Denos M, Mai X-M, Åsvold BO, Sørgjerd EP, Chen Y, Sun Y-Q. Vitamin D status and risk of type 2 diabetes in the Norwegian HUNT cohort study: does family history or genetic predisposition modify the association? **BMJ Open Diabetes Res Care**. 2021;9(1):e001948.

26. Cui JS, Hopper JL, Harrap SB. Antihypertensive treatments obscure familial contributions to blood pressure variation. **Hypertension**. 2003;41(2):207–210.

27. Tobin MD, Sheehan NA, Scurrah KJ, Burton PR. Adjusting for treatment effects in studies of quantitative traits: antihypertensive therapy and systolic blood pressure. **Stat Med**. 2005;24(19):2911–2935. 10.1002/sim.2165

28. Stenehjem JS, Hjerkind KV, Nilsen TI. Adiposity, physical activity, and risk of hypertension: prospective data from the population-based HUNT Study, Norway. **J Hum Hypertens**. 2018;32(4):278–286.

29. Bakris G, Ali W, Parati G. ACC/AHA Versus ESC/ESH on Hypertension Guidelines. **J Am Coll Cardiol**. 2019;73(23):3018–3026. doi:doi:10.1016/j.jacc.2019.03.507

30. Jiang L, Sun YQ, Brumpton BM, et al. Prolonged Sitting, Its Combination With Physical Inactivity and Incidence of Lung Cancer: Prospective Data From the HUNT Study. **Front Oncol**. 2019;9:101. doi:10.3389/fonc.2019.00101

31. Sun YQ, Langhammer A, Wu C, et al. Associations of serum 25-hydroxyvitamin D level with incidence of lung cancer and histologic types in Norwegian adults: a case-cohort analysis of the HUNT study. **Eur J Epidemiol**. Jan 2018;33(1):67–77. doi:10.1007/s10654-017-0324-1

32. Jiang L, Sun YQ, Langhammer A, et al. Asthma and asthma symptom control in relation to incidence of lung cancer in the HUNT study. **Sci Rep**. Feb 25 2021;11(1):4539. doi:10.1038/s41598-021-84012-3

33. Lawlor DA. Commentary: Two-sample Mendelian randomization: opportunities and challenges. **Int J Epidemiol**. 2016;45(3):908–915. doi:10.1093/ije/dyw127

34. Palmer TM, Sterne JA, Harbord RM, et al. Instrumental variable estimation of causal risk ratios and causal odds ratios in Mendelian randomization analyses. **Am J Epidemiol**. Jun 15 2011;173(12):1392–403. doi:10.1093/aje/kwr026

35. Lawlor DA, Harbord RM, Sterne JA, Timpson N, Davey Smith G. Mendelian randomization: using genes as instruments for making causal inferences in epidemiology. **Stat Med**. Apr 15 2008;27(8):1133–63. doi:10.1002/sim.3034

36. Davies NM, Holmes MV, Davey Smith G. Reading Mendelian randomisation studies: a guide, glossary, and checklist for clinicians. **BMJ**. Jul 12 2018;362:k601. doi:10.1136/bmj.k601

37. Bowden J, Davey Smith G, Burgess S. Mendelian randomization with invalid instruments: effect estimation and bias detection through Egger regression. **Int J Epidemiol**. 2015;44(2):512–525.

38. Bowden J, Davey Smith G, Haycock PC, Burgess S. Consistent Estimation in Mendelian Randomization with Some Invalid Instruments Using a Weighted Median Estimator. **Genet Epidemiol**. 2016;40(4):304–314. 10.1002/gepi.21965

39. Verbanck M, Chen CY, Neale B, Do R. Detection of widespread horizontal pleiotropy in causal relationships inferred from Mendelian randomization between complex traits and diseases. **Nat Genet**. May 2018;50(5):693–698. doi:10.1038/s41588-018-0099-7

40. Evangelou E, Warren HR, Mosen-Ansorena D, et al. Genetic analysis of over 1 million people identifies 535 new loci associated with blood pressure traits. **Nat Genet**. 2018/10/01 2018;50(10):1412-1425. doi:10.1038/s41588-018-0205-x

41. Kurki MI, Karjalainen J, Palta P, et al. FinnGen provides genetic insights from a well-phenotyped isolated population. **Nature**. 2023/01/01 2023;613(7944):508-518. doi:10.1038/s41586-022-05473-8

42. Burgess S. Violation of the constant genetic effect assumption can result in biased estimates for non-linear Mendelian randomization. **Humam Heredity**. 2023;88(1):79–90. doi:10.1159/000531659

43. Burgess S, Davies NM, Thompson SG. Instrumental variable analysis with a nonlinear exposure–outcome relationship. **Epidemiology (Cambridge**, **Mass**). 2014;25(6):877.

44. Mokhtari E, Hajhashemy Z, Saneei P. Serum Vitamin D Levels in Relation to Hypertension and Pre-hypertension in Adults: A Systematic Review and Dose–Response Meta-Analysis of Epidemiologic Studies. Systematic Review. **Front Nutr**. 2022-March-10 2022;9:829307. doi:10.3389/fnut.2022.829307

45. Paschou SA, Kosmopoulos M, Nikas IP, et al. The impact of obesity on the association between vitamin D deficiency and cardiovascular disease. **Nutrients**. 2019;11(10):2458.

46. Kunutsor SK, Burgess S, Munroe PB, Khan H. Vitamin D and high blood pressure: causal association or epiphenomenon? **Eur J Epidemiol**. Jan 2014;29(1):1–14. doi:10.1007/s10654-013-9874-z

47. Wang TJ, Zhang F, Richards JB, et al. Common genetic determinants of vitamin D insufficiency: a genome-wide association study. **Lancet**. Jul 17 2010;376(9736):180-8. doi:10.1016/s0140-6736(10)60588-0

48. Hamilton FW, Hughes DA, Spiller W, Tilling K, Smith GD. Non-linear mendelian randomization: evaluation of biases using negative controls with a focus on BMI and Vitamin D. **medRxiv**. 2023:08.21.23293658. doi:10.1101/2023.08.21.23293658

49. Langhammer A, Krokstad S, Romundstad P, Heggland J, Holmen J. The HUNT study: participation is associated with survival and depends on socioeconomic status, diseases and symptoms. **BMC Med Res Methodol**. Sep 14 2012;12:143. doi:10.1186/1471-2288-12-143

50. Burgess S, Butterworth A, Thompson SG. Mendelian randomization analysis with multiple genetic variants using summarized data. **Genet Epidemiol**. 2013;37(7):658–665.

## References

1. Sofianopoulou E KS, Afzal S, Jiang T, Gill D, Gundersen TE, et al. Estimating dose-response relationships for vitamin D with coronary heart disease, stroke, and all-cause mortality: observational and Mendelian randomisation analyses. **Lancet Diabetes Endocrinol** 2021; 9:837–846.

2. Jiang X, O’Reilly PF, Aschard H, Hsu YH, Richards JB, Dupuis J, et al. Genome-wide association study in 79,366 European-ancestry individuals informs the genetic architecture of 25-hydroxyvitamin D levels. **Nat Commun** 2018; 9:260.

3. Zhou A, Selvanayagam JB, Hyppönen E. Non-linear Mendelian randomization analyses support a role for vitamin D deficiency in cardiovascular disease risk. **Eur Heart J** 2022; 7;43(18):1731-1739.

4. Manousaki D, Mitchell R, Dudding T, Haworth S, Harroud A, Forgetta V, et al. Genome-wide Association Study for Vitamin D Levels Reveals 69 Independent Loci. **Am J Hum Genet** 2020; 106:327–337.

5. Revez JA, Lin T, Qiao Z, Xue A, Holtz Y, Zhu Z, et al. Genome-wide association study identifies 143 loci associated with 25 hydroxyvitamin D concentration. **Nat Commun** 2020; 11:1647.

6. Palmer TM, Sterne JA, Harbord RM, Lawlor DA, Sheehan NA, Meng S, et al. Instrumental variable estimation of causal risk ratios and causal odds ratios in Mendelian randomization analyses. **Am J Epidemiol** 2011; 173:1392–1403.

7. Sanderson E, Glymour MM, Holmes MV, Kang H, Morrison J, Munafò MR, et al. Mendelian randomization. Nat Rev Methods Primers 2022; 2:6.

8. Burgess S, Thompson SG. Use of allele scores as instrumental variables for Mendelian randomization. **Int J Epidemiol** 2013; 42:1134–1144.

9. Lawlor DA, Harbord RM, Sterne JA, Timpson N, Davey Smith G. Mendelian randomization: using genes as instruments for making causal inferences in epidemiology. **Stat Med** 2008; 27:1133–1163.

